# Multi-pathogen serosurveillance reveals correlated routine vaccination performance, waning tetanus immunity, and diphtheria boosting among children in Zambia

**DOI:** 10.64898/2026.06.01.26354612

**Authors:** Alyssa N. Sbarra, Simon Mutembo, Andrea Carcelen, Christine Prosperi, William J. Moss, Shaun Truelove, Amy K. Winter, Innocent C. Bwalya, Evans Betha, Lombe Kampamba, Elizabeth Kabeta, Gershom Chongwe, Amy Wesolowski, Saki Takahashi

## Abstract

**Background:** Vaccination coverage estimates and case-based surveillance have limitations in evaluating immunization programs. Serosurveillance offers a complementary approach by directly measuring population immunity. We assessed whether serologic analyses across multiple antigens (i.e., measles, diphtheria, tetanus) could provide additional insights into vaccination program performance.

**Methods:** We conducted a matched case-control study among children aged 2- to 10-years-old (n=1286) in Zambia using specimens from the 2016 ZAMPHIA survey. Using previously generated data on measles serostatus, measles seronegative children (i.e., “cases”) were matched to measles seropositive children (i.e., “controls”) on sex, age, HIV infection status, and province. Samples were tested for tetanus and diphtheria antitoxin IgG antibodies using commercial enzyme immunoassays. We estimated the odds of tetanus and diphtheria seropositivity by measles serostatus using conditional logistic regression and examined age-specific antibody dynamics.

**Results:** Measles seronegative children had 1.7-fold increased odds (95% credible interval [CrI]: 1.3-2.1) of being tetanus seronegative compared to measles seropositive children. Diphtheria serostatus had no significant association with measles serostatus (odds ratio: 1.3; 95% CrI: 0.9-1.7). Tetanus seroprevalence declined monotonically with age. However, diphtheria seroprevalence initially declined through 5 years of age, then increased again beginning at 6 years of age despite the lack of vaccine booster doses given after the primary series in infancy, potentially from asymptomatic or subclinical infections.

**Conclusions:** Serologic analyses revealed measles serostatus was positively associated with tetanus serostatus (where seropositivity arises only via vaccination and not infection), suggesting children who are measles seronegative are more likely to have missed DTP vaccination. We additionally found that measles serostatus was not associated with diphtheria serostatus, suggesting that antibody responses to diphtheria continue to boost beyond infancy when DTP vaccination is given. Our findings support consideration of DTP booster doses in Zambia to address waning tetanus immunity and further investigation of potential diphtheria carriage and transmission.

## Introduction

Despite the introduction of the Essential Programme on Immunization (EPI) over 50 years ago^1^, vaccine-preventable diseases (VPDs) still cause substantial morbidity and mortality, especially in low- and middle-income countries (LMICs).^2^ Although global vaccination coverage has increased, immunity gaps persist and countries require information on these gaps (e.g., by age, location) to better prioritize vaccination strategies, especially in settings with limited resources. Vaccination coverage estimates are typically derived from administrative data which have well-recognized biases, including errors in dose reporting, inaccurate target population estimates, and cross-border health use.^3–6^ Household surveys provide more representative information on vaccination history than administrative data, but are subject to infrequent implementation and sustainability challenges.^7,8^ Case-based surveillance of VPDs also has many limitations for evaluating vaccination program performance^9^, including challenges related to case ascertainment, asymptomatic transmission, case importations, and honeymoon or inter-outbreak or -epidemic periods. Importantly, as vaccination does not always result in protective immunity, neither coverage estimates nor case counts directly quantify population susceptibility, a key quantity of interest. Therefore, comprehensive evaluation of immunization program performance requires approaches that extend beyond vaccination coverage estimates or case-based surveillance alone.

The EPI program began with Bacillus Calmette-Guérin, diphtheria, tetanus, pertussis, polio, and measles vaccines, and has since expanded to include a broader group of childhood vaccinations. While the EPI program exists in counties as the cornerstone of national routine immunization programs, there are still vaccine- and antigen-specific considerations and challenges to address. For example, some VPDs (i.e., maternal and neonatal tetanus, and measles) are specifically targeted for elimination. Achieving and maintaining maternal and neonatal tetanus elimination will require sustained high population immunity and targeted interventions, particularly in LMICs.^10^ In Zambia, maternal and neonatal tetanus elimination has been reached, but because tetanus immunity wanes over time, maintaining elimination may require introduction of additional catch-up vaccination opportunities (e.g., booster doses).^10,11^ In addition, these strategies may be important for reaching populations not routinely captured through existing vaccination platforms (e.g., tetanus-toxoid or tetanus-diphtheria toxoid combination vaccines administered during antenatal care). For other VPDs, such as diphtheria and *S. pneumoniae*, surveillance and control programs have unique challenges.^12^ Asymptomatic carriage and subclinical infections can contribute to transmission, complicating the interpretation of reported incidence.^13^ For diphtheria particularly, a limited global supply of antitoxin further complicates clinical management and increases the need for ensuring high population immunity.^14^

Measles, also a target for elimination, causes substantial disease burden.^15,16^ Measles vaccination is first recommended at a later EPI touchpoint (i.e., 9-months-old in most LMICs^15^) compared to some other vaccinations (e.g., combined diphtheria-tetanus-pertussis [DTP], or DTP-HepB-Hib [pentavalent] vaccines; hereafter, DTP refers to DTP-containing-vaccines), which are recommended as early as 6 weeks of age in most LMICs.^10,17^ Dropout between vaccine doses and across vaccine series is well-documented, including within the primary 3-dose DTP series and from DTP to measles-containing vaccine (MCV), such that children reached with some vaccines may not be reached for other vaccines given later in life. First-dose DTP coverage is often used as a proxy metric for identifying “zero-dose” children^18^ (i.e., children who have never received any routine vaccinations). However, given the potential for DTP-to-MCV dropout, it is also important to consider immunity against other pathogens for which vaccination is administered later in childhood (i.e., measles) when considering the overall performance of a routine vaccination system. In Zambia, vaccination coverage estimates indicate that third-dose coverage of DTP and first-dose coverage of MCV were both high in 2016 (95% and 97%, respectively), with no evidence of DTP-to-MCV dropout; however, both have since declined at different rates, reaching 91% and 88% in 2024 respectively, underscoring the ongoing need to monitor dropout across the immunization cascade.^19,20^

Serology provides a complementary method to evaluate vaccination program performance by directly measuring antibody concentrations to determine serostatus. For most EPI VPDs, serostatus has been correlated as a marker of protection against infection or clinical disease.^21^ From individual serostatus, a proportion of persons susceptible in a population can be computed for each individual pathogen. Beyond single-antigen analyses, multi-pathogen serologic approaches show promise to comprehensively analyze cross-pathogen vulnerabilities and evaluate the performance of vaccination programs more holistically.^22^

Few studies to date have empirically examined correlations across multiple EPI antigens to assess routine immunization program performance and transmission dynamics simultaneously. This study evaluated whether serologic analyses across measles, diphtheria, and tetanus provide added insight into vaccination program performance and transmission. Specifically, this study assessed whether measles-seronegative children in Zambia were more likely to be diphtheria- and tetanus-seronegative using a matched case-control design and examined age-specific patterns of antitoxin IgG to infer waning and potential boosting.

## Methods

### Overview of primary study and biorepository

The 2016 Zambia Population-Based HIV Impact Assessment (ZAMPHIA) survey was a nationally representative, cross-sectional, multistage cluster household survey among 25383 residents aged 0- to 59-years-old.^23^ Parental consent was obtained and venous blood was collected from children aged 2- to 10-years-old in every other surveyed household. After primary ZAMPHIA testing, remaining plasma specimens were transported to and stored in a biorepository at the National Health Research and Training Institute (NHRTI), formerly Tropical Diseases Research Centre, in Ndola, Zambia.

A subset of specimens (n=9854) among children and adults aged 0- to 49-years-old from the biorepository were selected for additional testing to determine measles and rubella serostatus, the results of which have previously been published.^24^ Nationally-representative measles seroprevalence (i.e., proportion seropositive) for individuals aged 0- to 49-years-old was estimated to be 82.8% (95% confidence interval [CI]: 81.6 – 83.9%) and rubella seroprevalence was 74.9% (95% CI: 73.7 – 76.0%), with geographic heterogeneity.^24^ Notably, samples were collected prior to rubella vaccine introduction in Zambia in 2017.^25^

### Case-control sample selection and matching

To assess associations between measles serostatus with both diphtheria and tetanus serostatuses among children, we designed a matched case-control study where measles seronegative individuals were “cases” and measles seropositive individuals were “controls”. We restricted the analysis to children aged 2- to 10-years-old who were most likely to have completed their primary vaccination series for both DTP and MCV, but young enough to minimize the likelihood of extensive natural exposure to infection; further, the specimen type collected from children aged 0- to 1-year-old in the original ZAMPHIA survey was dried blood spots (DBS) rather than plasma, so we restricted testing to plasma only to circumvent potential variability introduced by specimen type.

We chose a matched case-control design to maximize statistical power for detecting associations between antigens given the relatively small number of measles seronegative (n=677) children in our target age range compared to combined positive (n=2217) and equivocal (n=745) results, which we collectively considered as seropositive (combined n=2962). Case-control matching also allowed for reducing potential confounding by demographic and geographic factors that could influence both vaccination history and serologic outcomes, and thus, further isolating associations between measles and diphtheria/tetanus serostatus. As such, because cases and controls were matched on age, the resulting odds ratios are inherently age-adjusted, removing the need for additional adjustment.

Samples with measles serostatus data available (n=3639) were selected for the matched case-control design in a two-staged matching approach in a 1:1 ratio without replacement. In the first stage (**Figure S1**), measles seropositives and seronegatives were identified for a stringent “Class 1” match (via MatchIt R package^26^) exactly by age in years, sex, HIV infection status, and province (i.e., first-administrative units), and by distance across districts (i.e., second-administrative units), such that districts closer in space were prioritized for matching. In the second phase, remaining unmatched samples were eligible for a less stringent “Class 2” matching which paired measles seropositive and seronegative samples by sex, HIV infection status and province, and by distance across age such that more similar ages were prioritized for matching. All identified Class 1 (n=1276 total samples, with n=638 matched pairs) and Class 2 (n=10 total samples, with n=5 matched pairs) matches were selected for diphtheria and tetanus serologic testing. In total, 1286 total samples from 643 matched pairs were selected for testing. Overall, this included 95.0% (643/677) of the total children aged 2- to 10-years-old who were measles seronegative in the original serosurvey.

### Laboratory methods and serostatus definitions

Serologic testing for diphtheria and tetanus was conducted at NHRTI from February to August 2024 using commercial indirect enzyme immunoassays (EIA; EUROIMMUN Anti-Diphtheria Toxoid IgG and VaccZyme^TM^ Anti-Tetanus Toxoid IgG). Plasma samples from the biorepository were processed as recommended by kit manufacturers. Antibody concentrations (IU/mL) were generated from optical density (OD) values of kit provided standards using a four-parameter logistic model.^27^

Serostatus definitions followed internationally recognized thresholds based on correlates of protection for both diphtheria and tetanus; samples with antitoxin IgG antibody concentrations above 0.1 IU/mL were considered seropositive for diphtheria^28^ and above 0.2 IU/mL were considered seropositive for tetanus.^29^ This tetanus threshold (0.2 IU/mL) is higher than is typically used by other assays to account for known non-specific binding of tetanus antitoxin IgG antibodies on EIAs.^29^ As per kit instructions, samples were classified with an “uncertain” serostatus for diphtheria if antitoxin IgG titers were between 0.01 and 0.1 IU/mL but to calculate binary serostatus, “uncertain” individuals were characterized as diphtheria seropositive.

### Statistical analysis

We computed diphtheria and tetanus antitoxin IgG seroprevalence by age, sex, and province across samples tested. Leveraging the matched case-control study design, we fit a Bayesian conditional logistic regression model (via R Stan^30^) to assess the odds of diphtheria and tetanus antitoxin IgG seropositivity among children who were measles seronegative compared to children who were measles seropositive. We additionally assessed differences in mean antitoxin IgG antibody concentration for both diphtheria and tetanus stratified by measles serostatus.

Cross-sectional serology data for VPDs across age groups have shown to be reasonable approximations of longitudinal serologic analyses of antibody levels as children age.^31^

Therefore, among all tested samples, we assessed correlations of diphtheria and tetanus antitoxin IgG seroprevalence and antibody concentrations by age.

To further examine age-specific patterns of diphtheria antitoxin IgG seroprevalence and antibody concentrations, we conducted additional statistical analyses. Modeling diphtheria serological data alone can be challenging given the complex nature of its pathogenesis, transmission characteristics, epidemiology, and related limitations in surveillance.^12^ Seropositivity for diphtheria antitoxin could be indicative of prior vaccination, infection, or asymptomatic carriage^13^, and the rate of boosting from exposure that results in carriage or infection remains uncharacterized, including differences in boosting following carriage versus infection.^12^ Given these limitations in interpreting diphtheria serostatus data, we chose to parameterize a simple decay and boosting model to capture changes in mean antitoxin IgG antibody concentrations by age to test against the hypothesis that there was no circulating *Corynebacterium diphtheriae* observed in the study cohort (i.e., as suggested by case-based surveillance reports^11^). We developed a Bayesian model in R Stan^30^ such that:

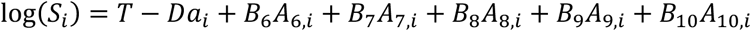

where *S_i_* is the diphtheria antitoxin IgG antibody concentration from individual *i*, *T* is the intercept, *D* is the decay rate, *a_i_* is the age (in years) of individual *i*. *B*_6_ is the age-specific boosting term for everyone aged 6-years-old and older (i.e., what boosting is expected in the sixth year of life). Therefore, *A*_6,*i*_ = 1 when *a_i_* ≥ 6. *B_X_* and *A_X_*_,*i*_ follow this same pattern for *X* values 7 through 10. We examined boosting specifically among children aged 6- to 10-years-old based on increasing mean antibody concentrations empirically observed among this age range. The model was fit to cross-sectional diphtheria antitoxin IgG antibody concentrations by age and convergence assessed via evaluations of R-hat. Mean annual decay rates across all ages and annual boosting rates for ages following the inflection point in seroprevalence were estimated from the joint posterior distribution.

All analyses were conducted in the R computing environment^32^ (version 4.3.2) and computer code is available online at https://github.com/alyssasbarra/diphtheria_tetanus_case_control.

## Results

We analyzed a final dataset of 1286 samples from individuals aged 2- to 10-years-old from 643 matched pairs (**Table 1**). Per the matched case-control design, 50% of individuals (i.e., 1 sample from each matched pair) was measles seronegative (“case”) and the other 50% was measles seropositive (“control”). 22 children were living with HIV, with 11 children living with HIV who were measles seropositive and 11 who were measles seronegative.

**Table 1.** Matched case-control study demographics. Measles seronegative individuals (i.e., cases) and seropositive individuals (i.e., controls) were matched on sex, province, age, and HIV status.

|  |  | Measles<br>seronegative<br>individuals | Measles<br>seropositive<br>individuals | Note |
| --- | --- | --- | --- | --- |
| Total (n, %) | – | 643 (50.0%) | 643 (50.0%) | – |
| Sex (n, %) | Male | 381 (50.0%) | 381 (50.0%) | (Class 1 and 2 required to<br>match exactly) |
|  | Female | 262 (50.0%) | 262 (50.0%) |  |
| Province (n, %) | Central | 39 (50.0%) | 39 (50.0%) | (Class 1 and 2 required to<br>match exactly) |
|  | Copperbelt | 75 (50.0%) | 75 (50.0%) |  |
|  | Eastern | 61 (50.0%) | 61 (50.0%) |  |
|  | Luapula | 69 (50.0%) | 69 (50.0%) |  |
|  | Lusaka | 98 (50.0%) | 98 (50.0%) |  |
|  | Muchinga | 88 (50.0%) | 88 (50.0%) |  |
|  | Northern | 37 (50.0%) | 37 (50.0%) |  |
|  | North-Western | 52 (50.0%) | 52 (50.0%) |  |
|  | Southern | 86 (50.0%) | 86 (50.0%) |  |
|  | Western | 38 (50.0%) | 38 (50.0%) |  |
| Age, years (n, %) | 2 | 43 (50.0%) | 43 (50.0%) | (Class 1 required match<br>exactly, Class 2 matched per<br>distance) |
|  | 3 | 73 (50.0%) | 73 (50.0%) |  |
|  | 4 | 84 (50.0%) | 84 (50.0%) |  |
|  | 5 | 92 (50.3%) | 91 (49.7%) |  |
|  | 6 | 65 (49.6%) | 66 (50.4%) |  |
|  | 7 | 71 (50.4%) | 70 (49.6%) |  |
|  | 8 | 72 (50.0%) | 72 (50.0%) |  |
|  | 9 | 65 (50.0%) | 65 (50.0%) |  |
|  | 10 | 78 (49.7%) | 79 (50.3%) |  |
| HIV status (n, %) | People not<br>living with HIV | 632 (50.0%) | 632 (50.0%) | (Class 1 and 2 required to<br>match exactly) |
|  | People living<br>with HIV | 11 (50.0%) | 11 (50.0%) |  |

### Tetanus seroprevalence

Overall, tetanus antitoxin IgG antibody seroprevalence was 52.2% (i.e., 671/1286 total samples). Among measles seropositive children, tetanus antitoxin IgG antibody seroprevalence was 58.0% (373/643 total samples) (**Table 2**) but only 46.3% (298/643 total samples) among children who were measles seronegative.

**Table 2.** Serologic results and odds ratios. Tetanus and diphtheria serologic results and corresponding odds ratios are included.

|  |  | Measles<br>seronegative<br>individuals | Measles<br>seropositive<br>individuals | Odds ratio (95% CI) |
| --- | --- | --- | --- | --- |
| Total (n, %) | – | 643 (50.0%) | 643 (50.0%) | – |
| Tetanus<br>serostatus (n, %) | Negative | 345 (56.1%) | 270 (43.9%) | 1.7 (1.3 – 2.1) |
|  | Positive | 298 (44.4%) | 373 (55.6%) |  |
| Diphtheria<br>serostatus (n, %) | Negative | 115 (55.0%) | 94 (45.0%) | 1.3 (0.9 – 1.7) |
|  | Positive | 528 (49.0%) | 549 (51.0%) |  |

Children who were measles seronegative had 1.7 (95% credible interval [CrI]: 1.3–2.1) times the odds of being tetanus seronegative compared to children who were measles seropositive. Mean tetanus antitoxin IgG antibody concentrations were significantly higher in measles seropositive individuals (mean = 0.516 IU/mL) compared to measles seronegative individuals (**Figure 2**; mean = 0.389 IU/mL; p < 0.001, Wilcoxon rank t-test). In a sensitivity analysis, we used a less stringent cutoff of 0.1 IU/mL rather than 0.2 IU/mL; tetanus seroprevalence was 82.9% (1066/1286 total samples) and children who were measles seronegative had 3.6 (95% CrI: 2.3–6.1) times the odds of being tetanus seronegative compared to children who were measles seropositive.

### Diphtheria seroprevalence

Diphtheria antitoxin IgG seroprevalence (positive and “uncertain” serostatuses) was 83.7% (**Table 2**). Among measles seropositive children, diphtheria antitoxin IgG seroprevalence was 85.4% (549/643 total samples) and 82.1% (528/643 total samples) among children who were measles seronegative. Children who were measles seronegative did not have an increased odds of being diphtheria seronegative compared to children who were measles seropositive (odds ratio: 1.3 [95% CrI: 0.9–1.7]).

However, mean diphtheria antitoxin IgG antibody concentrations were significantly higher in measles seropositive individuals (mean = 0.232 IU/mL) compared to measles seronegative individuals (**Figure 2**; mean = 0.175 IU/mL; p = 0.049, Wilcoxon rank t-test). In a sensitivity analysis, we only considered children with a positive diphtheria serostatus as seropositive (i.e., “uncertain” serostatuses were re-classified as seronegative); diphtheria seroprevalence was 34.6% (445/1286 total samples) and children who were measles seronegative still did not have an increased odds of being diphtheria seronegative compared to children who were measles seropositive (odds ratio: 1.2 [95% CrI: 0.9–1.7]).

### Age-specific serologic patterns

Diphtheria and tetanus antitoxin IgG seroprevalence changed markedly by age (**Figure 1**). The seroprevalence of tetanus antitoxin IgG decreased as age increased (Figure 1a). Similarly, the seroprevalence of diphtheria antitoxin IgG decreased as age increased (Figure 1b); however, this trend only persisted until age 5 years of age. For ages 6 years and above, the seroprevalence to diphtheria antitoxin IgG increased with age. Tetanus and diphtheria antitoxin IgG antibody concentrations were statistically significantly correlated among 2-year-olds, 3-year-olds and 4-year-olds (**Figure 3**; p<0.001, Pearson’s correlation). Generally, among children older than 5 years of age, tetanus and diphtheria antitoxin IgG antibody concentrations were no longer correlated (**Table S1**; p≥0.05, Pearson’s correlation).

**Figure 1.**
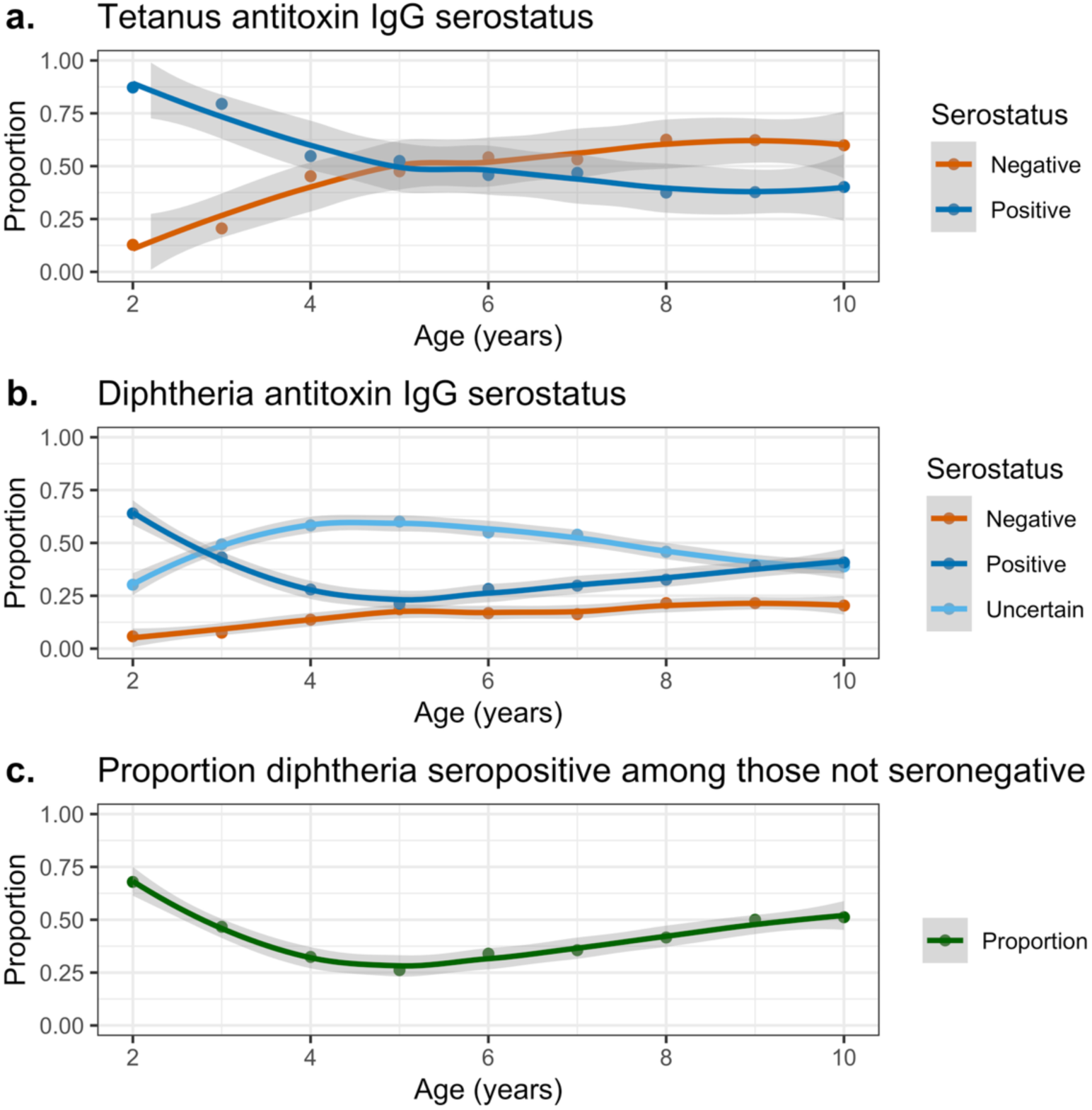
Tetanus antitoxin and diphtheria antitoxin IgG seroprevalence by age (in years). For tetanus (panel A), children were categorized as seropositive if tetanus antitoxin IgG antibody concentrations were greater than to 0.2 IU/mL and seronegative if less than 0.2 IU/mL. For diphtheria (panel B), children were categorized as seropositive if diphtheria antitoxin IgG antibody concentrations were greater than or equal to 0.1 IU/mL, uncertain if between 0.01 and 0.1 IU/mL, and seronegative if less than or equal to 0.01 IU/mL. Additionally (panel C), proportion of children diphtheria seropositive among those not seronegative (i.e., seropositive and uncertain) highlights the declining then increasing proportion seropositive among those with positive and uncertain serostatus.

**Figure 2.**
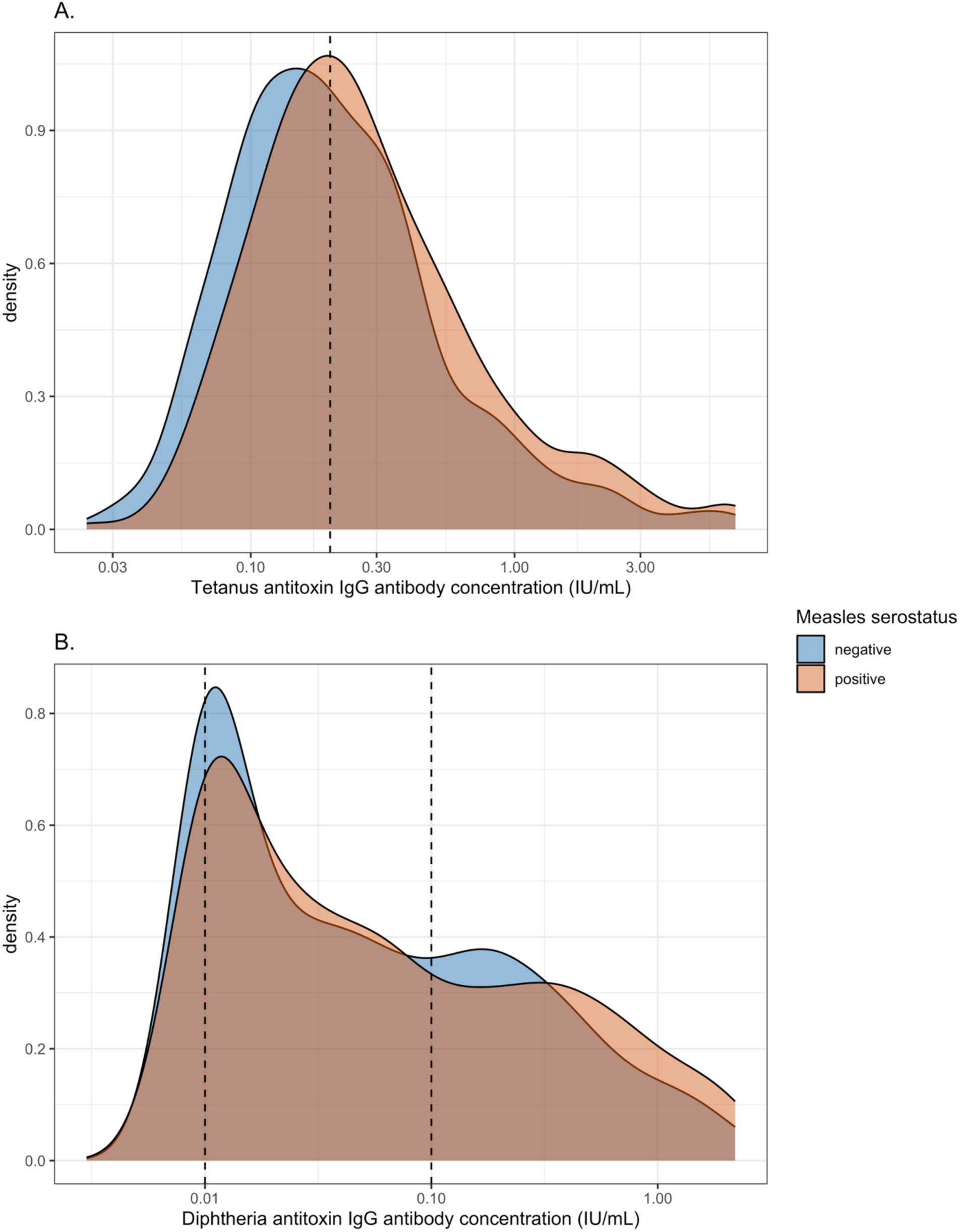
Tetanus and diphtheria antitoxin IgG antibody concentration by measles serostatus. Panel A for tetanus and panel B for diphtheria with vertical dashed lines at positive (and for diphtheria, uncertain) serostatus thresholds.

**Figure 3.**
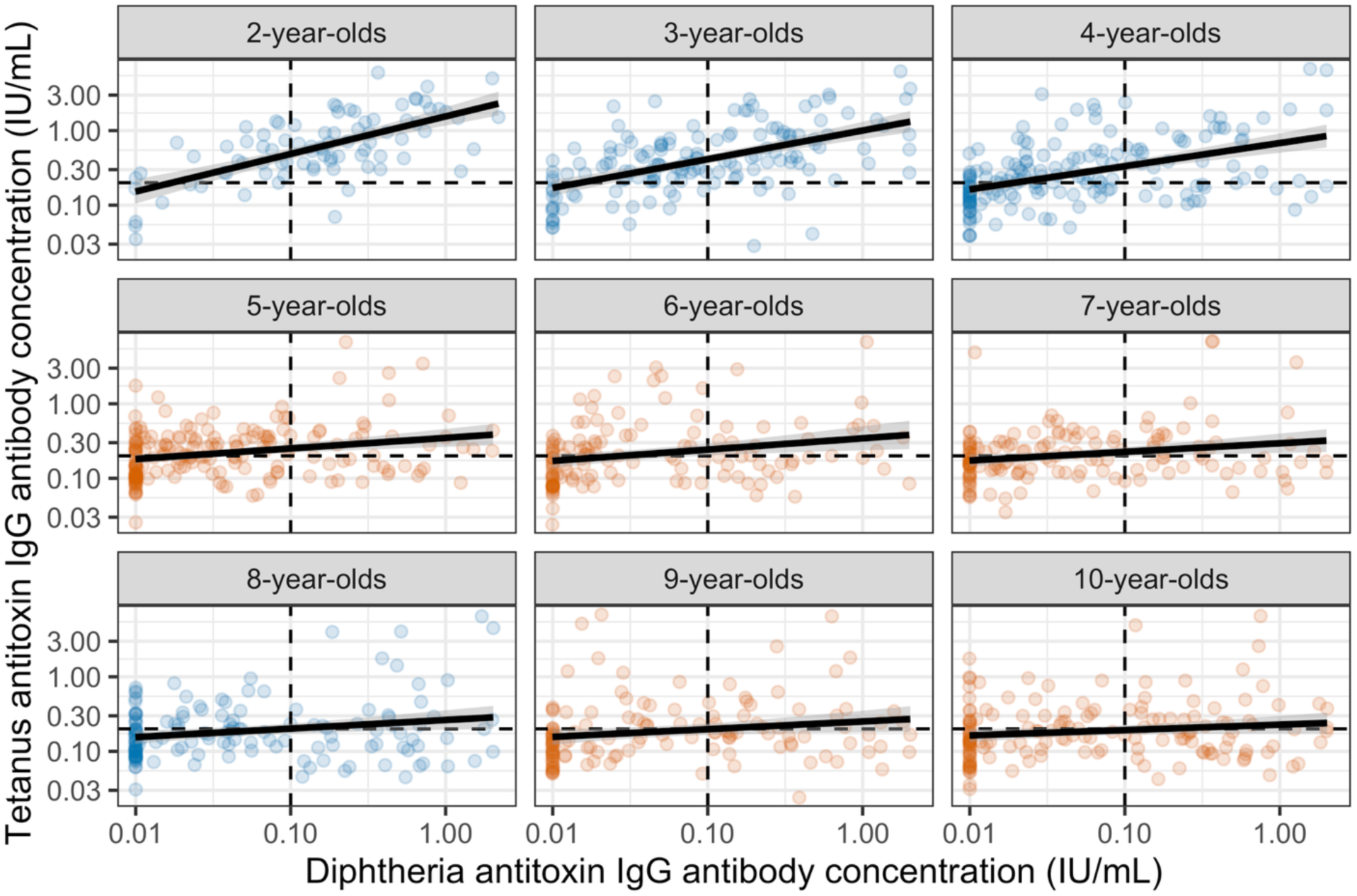
Tetanus and diphtheria antitoxin IgG antibody concentration by age (in years). Correlation significant when points (i.e., individuals) are displayed in blue (Table S1). Dotted lines correspond to seroprotective thresholds (0.1 IU/mL for diphtheria antitoxin and 0.2 IU/mL for tetanus antitoxin).

As with seroprevalence, the geometric mean antitoxin IgG antibody concentrations for diphtheria declined steeply in early childhood, crossing the protective threshold of 0.1 IU/mL by approximately ages 2- to 3-years-old. Antitoxin IgG antibody concentrations declined further until age 6 years when antibody concentrations modestly increased with age, consistent with possible boosting. Age-specific diphtheria antitoxin IgG antibody concentrations were modeled to estimate antibody dynamics and a counterfactual waning-only scenario (**Figure 4**).

**Figure 4.**
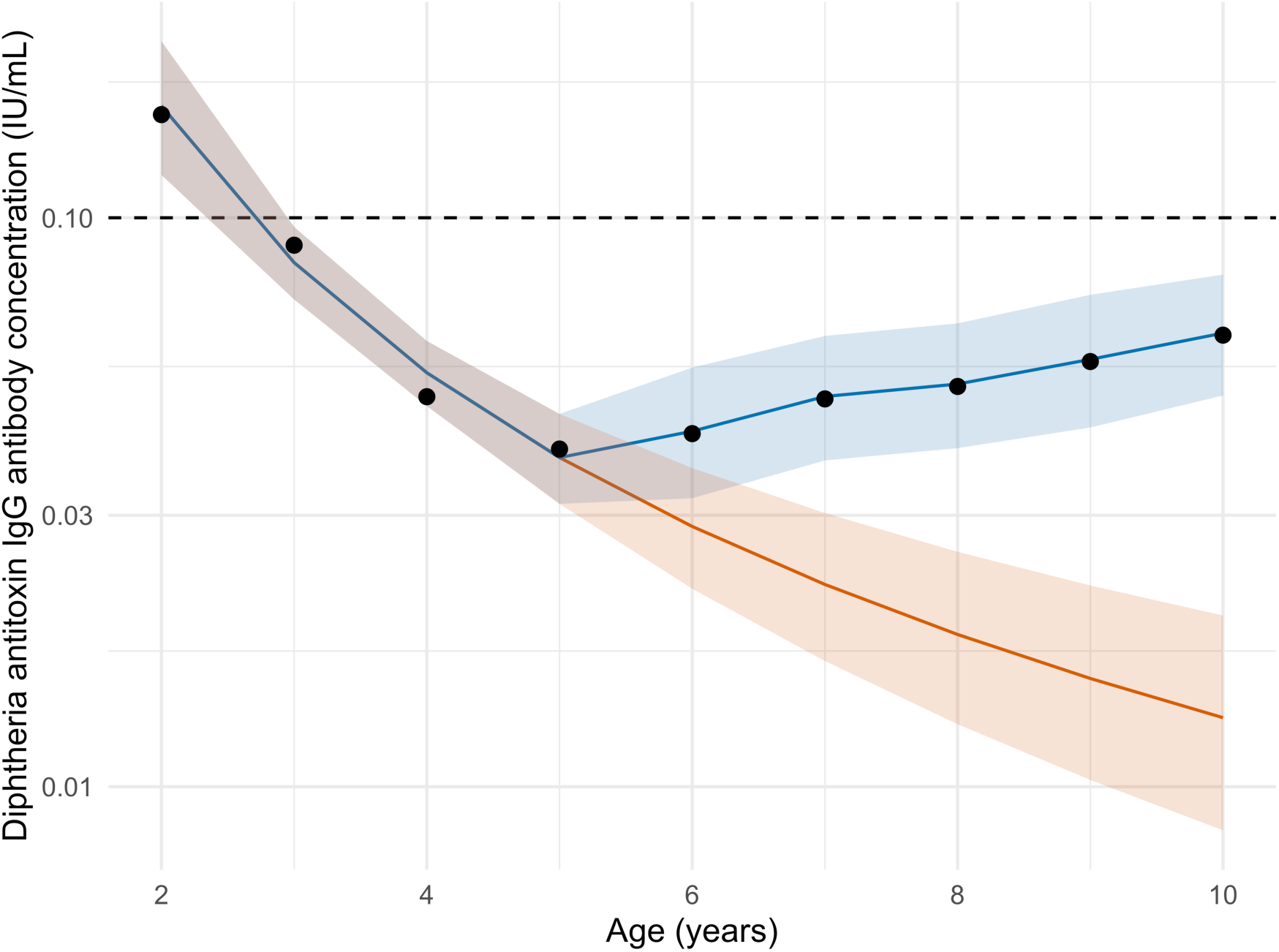
Mean diphtheria antitoxin IgG antibody concentration by age with waning and boosting and counterfactual (waning only) scenario. Dotted horizonal line at seroprotective threshold of 0.1 IU/mL. Each point is the mean diphtheria antitoxin IgG antibody concentration by age in years. Counterfactual waning only scenario shown in orange, and waning and boosting scenario shown in blue. 95% credible intervals shown in shaded ribbons.

The model (**Table S2**) identified a statistically significant age-related decline in the absence of boosting, with diphtheria antibody concentrations declining by 79% (95% credible interval: 68–86%) for each doubling of age. Each age beyond 5 years was estimated to have a positive cumulative boosting rate, with 6-year-olds (i.e., *T* − (*D* ∗ 6) + *B*_6_) having the lowest at 0.013 log of diphtheria antitoxin IgG antibody concentrations (95% CrI: 0.001–0.027) and 10-year-olds (i.e., *T* − (*D* ∗ 10) + *B*_6_ + *B*_$_ + *B*_%_ + *B*_&_ + *B*_’(_) having the highest at 0.049 log of diphtheria antitoxin IgG antibody concentrations (95% CrI: 0.034–0.067). Single age boosting was significant for 6-year-olds (*B*_6_= 0.382 log of diphtheria antitoxin IgG antibody concentrations [95% CrI: 0.034–0.738]) and 7-year-olds (*B*_$_= 0.381 log of diphtheria antitoxin IgG antibody concentrations [95% CrI: 0.005–0.751]). In contrast, the modeled counterfactual trajectory assumed that antibodies only wane (i.e., without any possibility of immune boosting). Under this counterfactual waning-only scenario, antitoxin IgG antibody concentrations continue to decline monotonically with age, reaching substantially lower levels by late childhood.

## Discussion

This study demonstrates that multi-antigen serology can provide valuable insights into vaccination program performance. Among Zambian children, measles serostatus was associated with tetanus but not diphtheria serostatus. Tetanus immunity waned monotonically over age, while diphtheria antibody levels initially waned over age followed by a subsequent increase, suggesting that immune responses are being boosted by some mechanism despite minimal reported cases. Analyzing measles, tetanus, and diphtheria antibodies together reveals patterns of vaccine access, immunity gaps, and potential pathogen exposure that administrative coverage data and case-based surveillance cannot detect on their own. We interpret these findings in the context of existing evidence and discuss implications for life-course immunization strategies, including booster policies.

Among Zambian children aged 2- to 10-years-old in year 2016, we found that measles seronegativity was significantly associated with tetanus antitoxin IgG seronegativity but, surprisingly, not with diphtheria antitoxin IgG seronegativity. The association between measles serostatus and tetanus antitoxin IgG serostatus likely reflects shared determinants of vaccine access and uptake; this was reflected in 55.8% of the children in our study with shared serostatus for both measles and tetanus. Measles seropositivity has previously been used as an indirect marker of immunization program reach, given its later administration and sensitivity to both missed routine services and drop-out.^33^ In this context the higher odds of tetanus seronegativity among measles seronegative children demonstrate potential shared structural determinants of vaccine access. Similar correlations across EPI antigens have been observed in other serosurveillance studies, reinforcing the value of cross-pathogen analyses for identifying systematically under-reached populations.^22^ Children who successfully received at least 1 dose of any MCV may represent those with better healthcare access or whose caregivers are more engaged with the immunization schedule, which in turn makes these children more likely to have also received at least one dose of the earlier DTP series. In our study, we identified nearly twice the odds of tetanus antitoxin IgG seropositivity among measles seropositive children as compared to measles seronegative children, supporting this hypothesis. A negative association between measles and tetanus serostatus (OR < 1) may have suggested either substantial dropout of children between the DTP and measles vaccine series, or that children were not receiving tetanus vaccination despite being vaccinated for measles, which could happen if routine vaccinations were inaccessible but children received measles vaccine during a campaign (as campaigns for DTP do not occur). Similarly, we found that measles seropositive children had significantly higher mean antitoxin IgG antibody concentrations for both tetanus and diphtheria, suggesting that children reached by the immunization program at the later MCV touchpoint may have had more consistent engagement with vaccination services overall, if receiving all 3 recommended DTP primary series doses leads to higher antibody levels.

Beyond access related patterns, our study contributes to the growing body of evidence documenting waning tetanus immunity following primary infant vaccination series. Previous serological studies from diverse settings have shown that tetanus antitoxin levels decline steadily in the absence of booster doses, often reaching sub-protective levels by late childhood or adolescence.^10^ Our observation that tetanus antitoxin IgG seroprevalence fell below 50% by age 6 years is consistent with these reports and highlights the limitations of relying on high DTP coverage alone to infer durable population-level immunity. Notably these immunity gaps were observed even among children likely to have accessed routine immunization services, underscoring that waning immunity represents a programmatic challenge distinct from coverage shortfalls.

In contrast to tetanus, diphtheria antitoxin IgG exhibited a non-monotonic age pattern, with declining antibody levels in early childhood followed by increasing concentration among older children. This divergence is particularly informative given that both antigens are administered together via a combination vaccine, such that initial immunization exposures and serostatus should be similar. The significant correlation between tetanus and diphtheria antitoxin IgG antibody concentrations among young children (i.e., 2- to 4-year-olds) that was not present among older children further supports the interpretation that some additional exposure source is boosting diphtheria immunity but not tetanus immunity in older children. Towards this point, our modeling analyses estimated significant immune boosting beginning at age 6 years, coinciding with the age of school entry in Zambia, and this cumulative boosting effect increased with age. This pattern is consistent with potential asymptomatic carriage or subclinical infection that boosts antibody levels as children enter environments with increased social mixing.^12,34,35^ Notably, this serologic signal of diphtheria circulation stands in stark contrast to a lack of cases reported via case-based surveillance data (i.e., only two cases reported in Zambia since 2008^11^).

Given the rapid waning of tetanus immunity as well as the increases in diphtheria antibody concentrations in older children, these data support consideration of routine DTP booster dose introduction at or prior to school entry.^36^ Such boosters could be delivered at opportune contact points (e.g., school entry checks or school-based platforms^37^ or integrated with other health services or immunization program touchpoints^38^). The observation that nearly half of measles seropositive children (i.e., those presumably better connected to health services, assuming that measles seropositivity is a result of vaccination and not infection) were nonetheless tetanus seronegative suggests that waning immunity affects even populations with high vaccination coverage, and achieving and maintaining high population immunity may not be addressed solely through improved primary series coverage.

Beyond this specific analysis, our findings highlight broader opportunities for multi-pathogen serosurveillance as a tool for vaccination program evaluation and other public health applications by directly measuring population immunity^39,40^, regardless of availability or completeness of data on vaccination history. Analyses of antibody responses across antigens, as demonstrated here, can reveal patterns of correlated susceptibility that identify populations at heightened susceptibility to multiple VPDs, as well as divergent patterns that may signal potential pathogen transmission. To facilitate comparisons like these, future serosurveillance initiatives should consider multi-antigen panels that enable such cross-pathogen comparisons in a scalable, cost-effective manner and include collection of individual-level vaccination history for additional validation.

This study has several limitations. First, measles seropositivity in our sample could reflect either successful vaccination and/or natural infection, and we cannot distinguish between these sources of immunity. This limits our true ability to definitively attribute the identified association between measles and tetanus serostatus to vaccine access patterns rather than other unmeasured confounders such as exposure to measles infection. While not definitive, the last large measles outbreak in Zambia occurred from 2010 to 2011, so children in the 2016 ZAMPHIA survey aged older than 6 years are more likely than children aged less than 6 years to have measles antibodies from infection rather than vaccination. Additionally, our matched case-control design, while efficient for assessing associations between serostatus to multiple

VPDs, produced a non-representative sample in which, by design, half of the children were measles seronegative. Therefore, seroprevalence estimates from this sample should <u>not</u> be interpreted as representative of the population. Also, our use of EIAs for serologic testing may have limited our ability to detect low antibody concentrations, particularly for tetanus antitoxin IgG^29^, as more sensitive assays (e.g., Luminex multiplex immunoassays) have demonstrated superior performance in detecting low antibody levels. Finally, diphtheria presents unique challenges for serologic modeling due to incompletely characterized antibody kinetics following vaccination, natural infection, and asymptomatic carriage or subclinical infection. The amount of antibody boosting from exposure events that may or may not result in carriage or infection remains poorly understood. Additionally, we lacked data on antibiotic use patterns, which could affect diphtheria natural history and transmission dynamics, and subsequently, antibody responses. We were unable to fit more complex dynamic transmission models for diphtheria given these multiple unknowns, and we could not validate our findings against independent measures of asymptomatic transmission due to current lack of data availability. Nevertheless, the observed pattern of age-specific boosting is most parsimoniously explained by some form of natural exposure, and these findings invite further investigation into diphtheria transmission dynamics in settings with high vaccination coverage but limited surveillance capacity, with specific focus on subclinical infection and transmission.

Our findings support the recommendation that vaccination programs should consider additional DTP booster doses beyond the primary infant series. Additionally, serologic evidence suggests that asymptomatic diphtheria infections may have or be occurring. Given the limited global supply of diphtheria antitoxin and the potential for outbreaks when population-level immunity wanes, further investigation into diphtheria transmission dynamics in Zambia and comparatively in other settings – with and without routine DTP booster doses – is warranted. Overall, serologic analyses across VPD antigens can enhance understanding of vaccination program performance and disease transmission in ways that complement traditional coverage and surveillance metrics and be used to mitigate preventable morbidity and mortality across VPDs.

## Data availability statement

The de-identified data set used for this analysis has been uploaded to (https://github.com/alyssasbarra/diphtheria_tetanus_case_control).

