## Supplementary Data for "Multi-pathogen serosurveillance reveals correlated routine vaccination performance, waning tetanus immunity, and diphtheria boosting among children in Zambia"

**Figure S1. Case-control matching process.**
Class 1 and 2 matching process for case-control design.

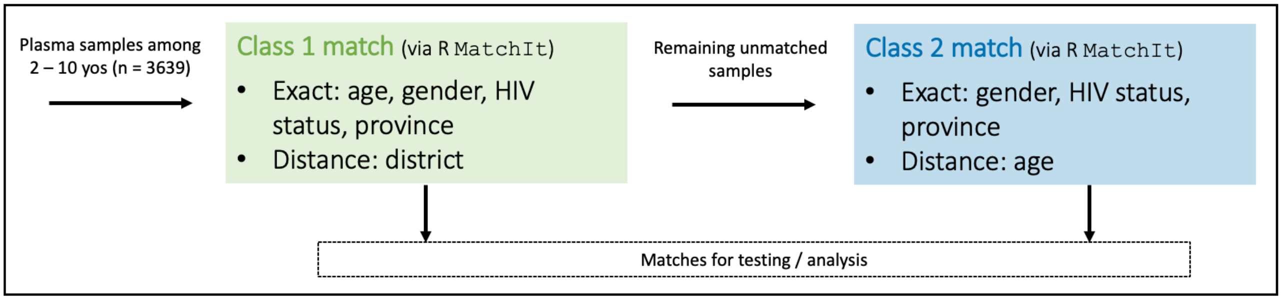

**Table S1. Anti-diphtheria and anti-tetanus toxin IgG antibody concentration (IU/mL) correlation by age.**

| Age (years) | Correlation | P-value |
|---|---|---|
| 2 | 0.494 | <0.001 |
| 3 | 0.499 | <0.001 |
| 4 | 0.504 | <0.001 |
| 5 | 0.110 | 0.137 |
| 6 | 0.166 | 0.059 |
| 7 | 0.111 | 0.192 |
| 8 | 0.398 | <0.001 |
| 9 | 0.021 | 0.816 |
| 10 | 0.085 | 0.291 |

**Table S2. Estimated diphtheria boosting model parameter values.**

| Parameter | Median | 95% Credible Interval |
|---|---|---|
| $T$ | -0.788 | -1.326 – -0.249 |
| $D$ | -1.548 | -1.956 – -1.138 |
| $B_6$ | 0.382 | 0.034 – 0.738 |
| $B_7$ | 0.381 | 0.005 – 0.751 |
| $B_8$ | 0.258 | -0.106 – 0.622 |
| $B_9$ | 0.280 | -0.091 – 0.657 |
| $B_{10}$ | 0.272 | -0.092 – 0.636 |
